# Differentiation of Medulloblastoma Molecular Subtypes Using Multiparametric MRI and Texture Analysis

**DOI:** 10.1101/2025.08.21.25333936

**Authors:** Bulent Aslan, Erhan Biyikli, Tahsin Aybal, Gulnur Tokuc, Suheyla Bozkurt, Nuri Cagatay Cimsit

## Abstract

**Background:** Medulloblastoma is the most common pediatric malignant brain tumor in the population, with molecular subtypes that differ in prognosis and therapeutic response. Identifying these subtypes before surgery is important for tailoring management.

**Objective:** This study aimed to distinguish medulloblastoma molecular subtypes using a combination of conventional MRI features and MRI-based texture analysis.

**Materials and methods:** We retrospectively analyzed 58 patients with preoperative MRI and histopathologic confirmation of medulloblastoma. Cases were classified into SHH pathway–activated or Group 3/4 subtypes. Morphologic MRI characteristics, apparent diffusion coefficient (ADC) ratios, and texture analysis parameters were compared between the groups.

**Results:** Of the 58 patients, 55.2% had SHH pathway–activated tumors. Morphological features, including location out of the midline or in the cerebellar hemisphere (p<0.001), peri-tumoral edema (p=0.041), macrocysts (p=0.001), nodular involvement/lobulation (p=0.002), and heterogeneous contrast enhancement (p=0.002) were more common in SHH tumors. ADC measurements showed that the solid tumor-to-thalamus ratio was significantly lower in SHH tumors (p<0.001), with a threshold of 0.855 providing 82.1% sensitivity and 92.3% specificity. As for texture analysis parameters, kurtosis (p=0.023), SumOfSqs (p=0.022) and 01-10-50-90% percentile (p=0.011; p=0.001; p=0.006; and p=0.013 respectively) values obtained from ADC images and kurtosis (p=0.041), SumOfSqs (p=0.005), SumVarnc (p=0.014), SumEntrp (p=0.032) values obtained from T1W images were statistically significant in differentiating SHH and group 3/ group 4 medulloblastoma.

**Conclusion:** Integrating MRI morphological features, ADC-based measurements, and texture analysis provides complementary information for non-invasive differentiation of medulloblastoma molecular subtypes.

## INTRODUCTION

Medulloblastoma (MB) represents the most common embryonal tumor of the central nervous system in children and is a major contributor to morbidity and mortality in this age group [1]. Once regarded as a single pathological entity, advances in molecular profiling have revealed that MB comprises at least four biologically distinct subgroups; Sonic Hedgehog (SHH), Wingless (WNT), Group 3, and Group 4, each with specific developmental origins, prognoses, and treatment implications [2–4].

Molecular classification is usually performed on surgical or biopsy specimens. In contrast to high-grade gliomas, MBs tend to display consistent genetic features throughout the lesion, allowing small samples to accurately determine the molecular subtype [5]. Nevertheless, tissue acquisition is invasive and carries procedural risks. While molecular subtyping can provide valuable prognostic and therapeutic guidance, routine implementation in clinical workflows has been hindered by cost, limited access to specialized testing, and the need for advanced laboratory techniques [4].

Although radiomics applications in MB are still relatively limited, numerous MRI-based studies have examined qualitative imaging features to predict molecular grouping. Tumor location and enhancement patterns are among the most consistently reported distinguishing features [6–11]. For example,

- Group 3 and Group 4 tumors frequently arise in the midline,
- SHH tumors more often originate within the cerebellar hemispheres, and
- WNT tumors may occur in the midline, cerebellar peduncle, or cerebellopontine angle cisterns [6, 8, 11].

In addition, a lack of contrast enhancement on MRI strongly suggests Group 4 disease, whereas marked enhancement in non-WNT/SHH tumors has been linked to less favorable outcomes [11].

Texture analysis is an advanced image-processing technique that quantifies tissue heterogeneity by extracting numerical features from specific regions of interest [12, 13]. Using statistical descriptors of image pixel intensity distributions, this method can objectively characterize tumor microstructure beyond what is visible to the human eye [14]. Texture features are generally organized by complexity:

- **First-order metrics** (e.g., mean intensity, standard deviation, entropy, kurtosis, skewness) summarize the overall statistical distribution of voxel intensities.
- **Second-order metrics** describe the spatial relationships between voxel intensities, capturing patterns such as coarseness or regularity.
- **Higher-order metrics** evaluate more complex spatial relationships, often requiring additional image transformations or filtering [15].

Developing reliable, non-invasive imaging biomarkers capable of predicting MB subtypes would enhance prognostication and aid in treatment planning without the need for surgical sampling. This study investigates whether a combination of multiparametric MRI features including conventional imaging findings, ADC measurements, and advanced texture analysis can be used to differentiate molecular subtypes of medulloblastoma.

## MATERIALS AND METHODS

This retrospective study was conducted with approval from our institutional review board. Owing to its retrospective design, the requirement for informed consent was waived.

### Patient Selection

We reviewed medical records for patients with histologically confirmed medulloblastoma who underwent posterior fossa surgery or biopsy at our tertiary referral center between January 2012 and August 2020. Ninety-one eligible cases were initially identified. We excluded 33 patients due to:

- absence of molecular subtype classification,
- suboptimal MRI quality from motion or technical artifacts, or
- Lack of preoperative MRI performed at our institution.

Of the excluded cases, only two had confirmed WNT-subtype medulloblastomas. The final cohort consisted of 58 patients with available preoperative MRI and confirmed SHH or Group 3/4 molecular classification. Patients were stratified into SHH-activated or Group 3/4 subgroups, and their MRI findings, ADC ratios, and tissue analysis metrics were compared.

### MRI Acquisition Protocol

Examinations were performed on either 1.5 T (Avanto) or 3 T (Verio) Siemens scanners using standardized brain MRI protocols. These included:

- **T1-weighted spin-echo**: TR 450–515 ms, TE 10–15 ms, flip angle 90°, FOV 200–260 mm
- **T2-weighted spin-echo**: TR 4200–4650 ms, TE 90–100 ms, flip angle 150°, FOV 200–260 mm
- **T2-FLAIR**: TR 7000–8000 ms, TE 84–100 ms, flip angle 150°, FOV 200–260 mm
- **Axial diffusion-weighted imaging (DWI)** with b-values of 0 and 1000 s/mm^2^, from which ADC maps were generated.

Slice thickness ranged from 3–4 mm. Post-contrast T1-weighted images were acquired using the same parameters as pre-contrast T1, with added fat suppression and sagittal acquisitions. The presence of hemorrhage or calcification was assessed in 31 patients using susceptibility-weighted imaging (SWI) or head CT, when available. Four patients lacked DWI, and three had no post-contrast T1-weighted imaging.

### Lesion Evaluation

Brain MRI images were initially evaluated blind to clinical information and postoperative histopathological diagnoses. On brain MRI, tumor localization, contrast enhancement pattern (heterogenous vs homogenous or minimal/non-enhancing), margins, T2 signal relative to cerebral gray matter, the presence of vasogenic edema, hemorrhage/calcification, microcysts (<1 cm)/macrocysts (>1 cm), nodularity/lobulation, and the presence of intracranial/spinal seeding or hydrocephalus were assessed.

ADC values of the lesions were measured using the PACS system. Measurements were made by drawing manual ROIs (regions of interest) to cover the solid part of the lesion. Additionally, the ADC value of the left thalamus was measured similarly in each patient for a reference value. The ration of mean ADC values of the solid part of the lesion and the left thalamus were calculated. Finally, the images of 58 patients (one ADC map, one T2-weighted image, and one post-contrast T1-weighted image per patient) were analyzed for texture analysis.

Texture analysis and histogram evaluations were performed using the MaZda program (MaZda 4.60, The Technical University of Lodz, Institute of Electronics, Poland). Each lesion was manually marked with an ROI to include the entire solid part of the lesion on a single slice (Figure 1). As a result, texture analysis data were extracted for three images of each lesion (one ADC map, one T2-weighted image, and one post-contrast T1-weighted image).

**Figure 1.**
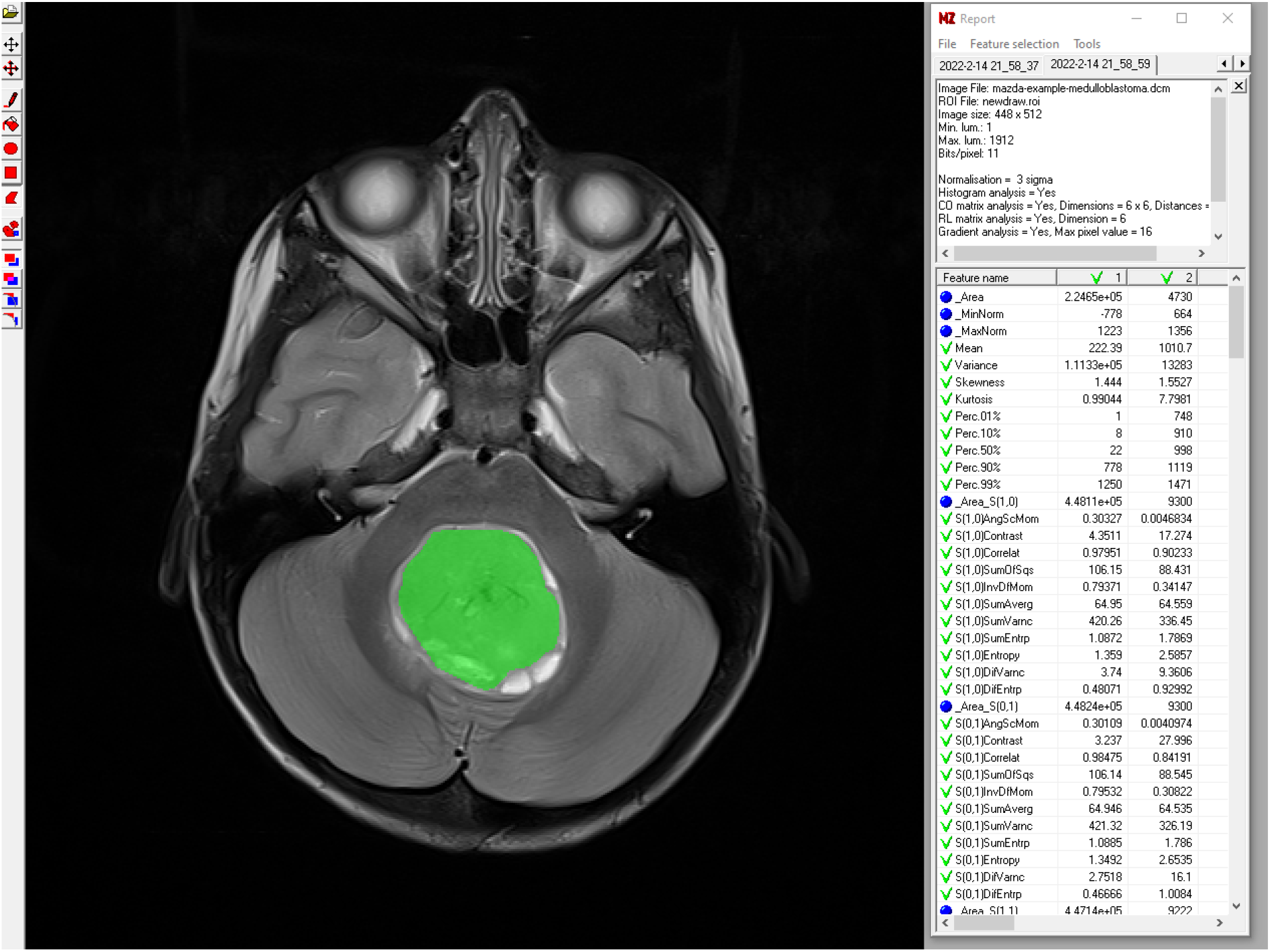
ROI segmentation of medulloblastomas for texture analysis

### Statistical Analysis

Data analyzes were performed using SPSS version 22 (IBM Corp., Armonk, NY, USA). Continuous variables were summarized as mean ± standard deviation or median with range, as appropriate. Normality was assessed with the Kolmogorov–Smirnov test. Group comparisons used the t-test for normally distributed data or the Mann–Whitney U test for non-normal distributions. ROC curve analyzes were conducted to determine the diagnostic performance of significant parameters. A p-value < 0.05 was considered statistically significant.

## RESULTS

The study included 58 patients with a mean age of 9.62 ± 8.25 years (range: 1 month–35.75 years). Males accounted for 58.6% of the cohort, and 55.2% of tumors were classified as SHH pathway–activated, with the remainder falling into Group 3/4.

### ADC Measurements

Table 1 summarizes ADC parameters, including the tumor solid-to-thalamus ratio (ratio of mean ADC in the tumor’s solid component to that of the normal left thalamus) and extracted texture metrics. Significant differences in ADC-based measurements were observed between molecular subtypes. Compared with Group 3/4 tumors, SHH pathway– activated tumors demonstrated:

**Table 1:**
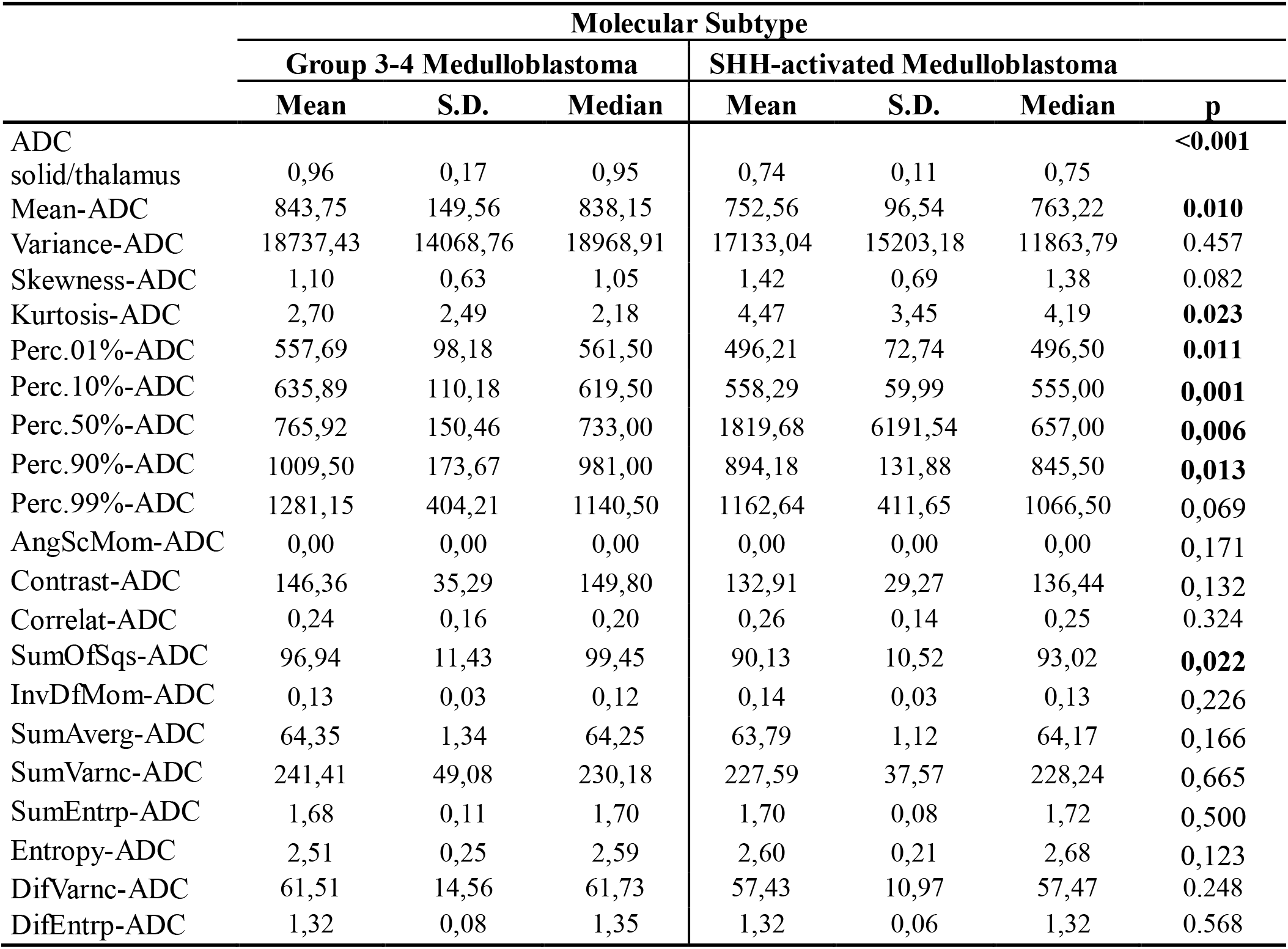
Comparison of MRI ADC measurements according to molecular subtypes.

- lower values for the solid-to-thalamus ratio, mean ADC, Percentiles 01%, 10%, 90%, and SumOfSqs, and
- higher values for ADC kurtosis and Percentile 50% (all p < 0.05).

The ROC analysis showed that the ADC solid tumor-to-thalamus ratio yielded the highest area under the curve (AUC = 0.891), indicating strong discriminatory capability (Figures 2 and 3). A threshold of 0.855 provided 82.1% sensitivity and 92.3% specificity, with corresponding positive and negative likelihood ratios of 10.6 and 0.19, respectively.

**Figure 2.**
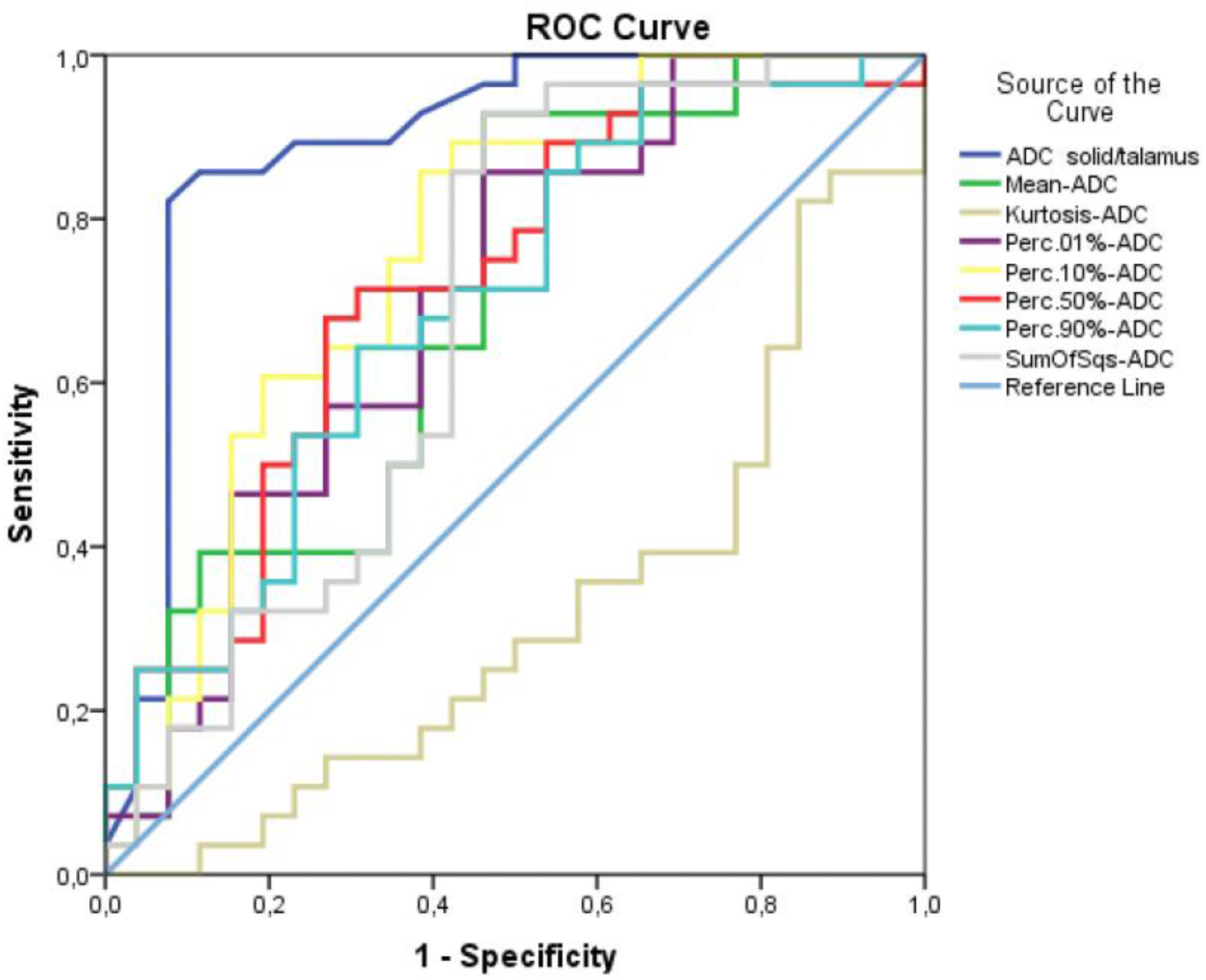
ROC analysis for ADC measurements

**Figure 3.**
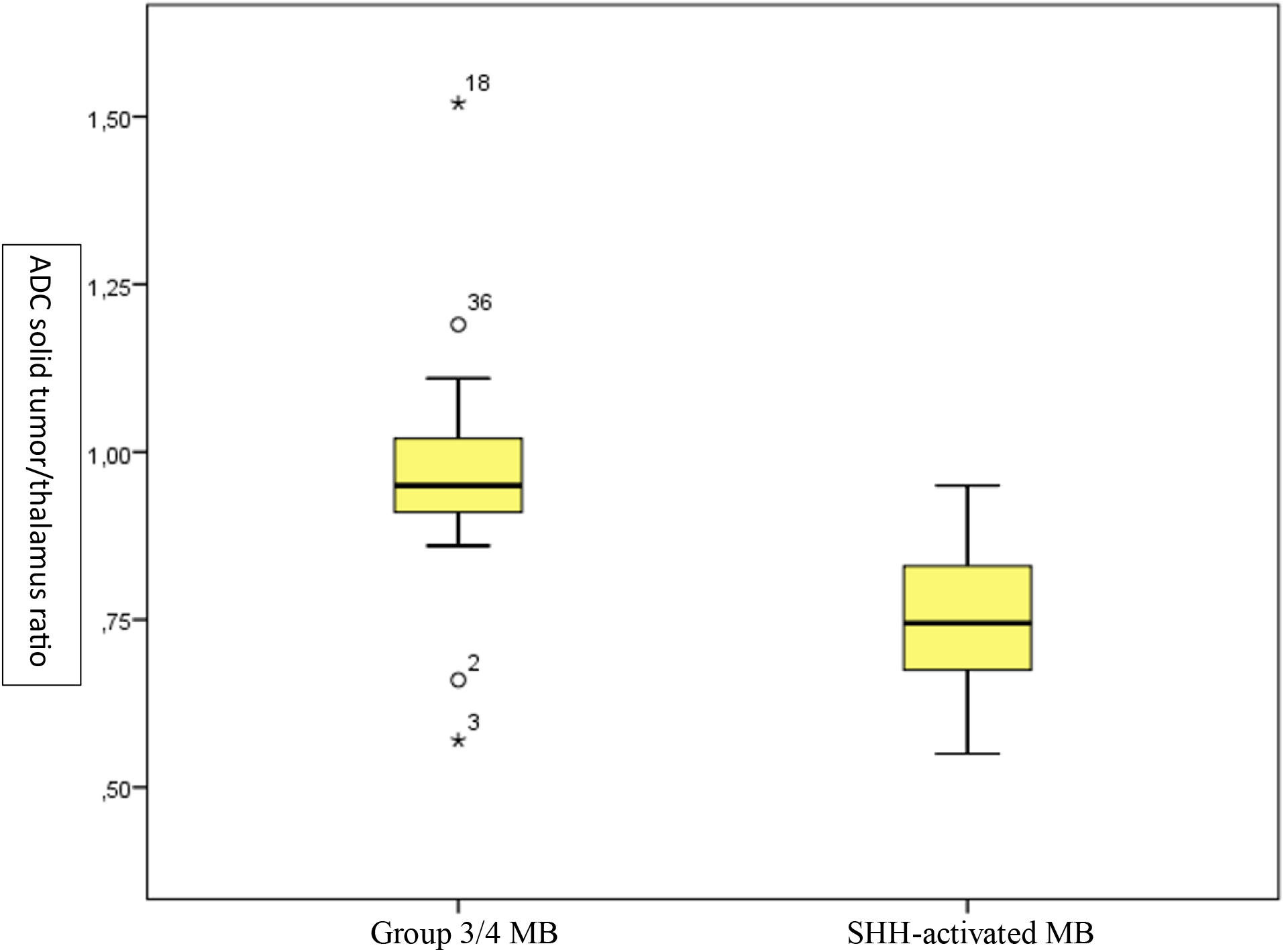
ADC solid tumor/thalamus ratio values according to molecular subtype (box-plot graph)

Table-1: Comparison of MRI ADC measurements according to molecular subtypes

### Texture Analysis – T1-weighted Features

From T1-weighted images, kurtosis, SumOfSqs, SumVarnc, and SumEntrp values differed significantly between subgroups. Lower kurtosis values tended to favor the SHH subtype, whereas the remaining three parameters were higher in SHH tumors (Table 2). Of these, SumOfSqs had the largest AUC in ROC analysis (cut-off = 107.06), achieving 62.1% sensitivity and 84.6% specificity (Figure 4).

**Table 2:** Comparison of MRI T1-weighted sequence measurements according to molecular subtypes.

|  | Molecular Subtype |  |  |  |  |  |  |
| --- | --- | --- | --- | --- | --- | --- | --- |
|  | Group 3-4 Medulloblastoma |  |  | SHH-activated Medulloblastoma |  |  |  |
|  | Mean | S.D. | Median | Mean | S.D. | Median | p |
| Mean-T1 | 430,87 | 251,18 | 354,90 | 1626,41 | 6254,52 | 380,85 | 0,522 |
| Variance-T1 | 5104,53 | 4923,56 | 3470,40 | 10205,11 | 14241,63 | 4989,36 | 0,134 |
| Skewness-T1 | 0,30 | 0,92 | 0,08 | 0,00 | 0,94 | -0,12 | 0,125 |
| Kurtosis-T1 | 1,49 | 2,53 | 0,68 | 1,55 | 6,93 | -0,12 | **0,041** |
| Perc.01%-T1 | 299,62 | 183,72 | 239,50 | 287,86 | 192,27 | 229,00 | 0,800 |
| Perc.10%-T1 | 360,77 | 220,75 | 272,00 | 365,38 | 226,35 | 310,00 | 0,794 |
| Perc.50%-T1 | 427,62 | 255,93 | 359,50 | 465,10 | 279,00 | 381,00 | 0,631 |
| Perc.90%-T1 | 506,50 | 281,73 | 438,00 | 549,35 | 314,10 | 440,00 | 0,853 |
| Perc.99%-T1 | 579,65 | 303,64 | 544,50 | 606,07 | 333,91 | 497,00 | 0,926 |
| AngScMom-T1 | 0,00 | 0,00 | 0,00 | 0,00 | 0,00 | 0,00 | 0,428 |
| Contrast-T1 | 85,55 | 44,02 | 80,32 | 69,62 | 30,56 | 70,29 | 0.122 |
| Correlat-T1 | 0,57 | 0,22 | 0,57 | 0,67 | 0,15 | 0,67 | 0.055 |
| SumOfSqs-T1 | 100,55 | 7,86 | 102,05 | 105,30 | 8,50 | 107,78 | **0,005** |
| InvDfMom-T1 | 0,20 | 0,09 | 0,18 | 0,22 | 0,08 | 0,19 | 0,238 |
| SumAverg-T1 | 65,09 | 1,18 | 65,26 | 65,34 | 0,84 | 65,32 | 0,601 |
| SumVarnc-T1 | 316,65 | 55,43 | 320,03 | 351,56 | 46,02 | 355,06 | **0,014** |
| SumEntrp-T1 | 1,80 | 0,08 | 1,82 | 1,83 | 0,04 | 1,85 | **0,032** |
| Entropy-T1 | 2,76 | 0,22 | 2,81 | 2,82 | 0,13 | 2,85 | 0,281 |
| DifVarnc-T1 | 36,49 | 16,50 | 34,57 | 31,01 | 12,54 | 31,66 | 0.169 |
| DifEntrp-T1 | 1,20 | 0,15 | 1,21 | 1,16 | 0,14 | 1,17 | 0.218 |

**Figure 4.**
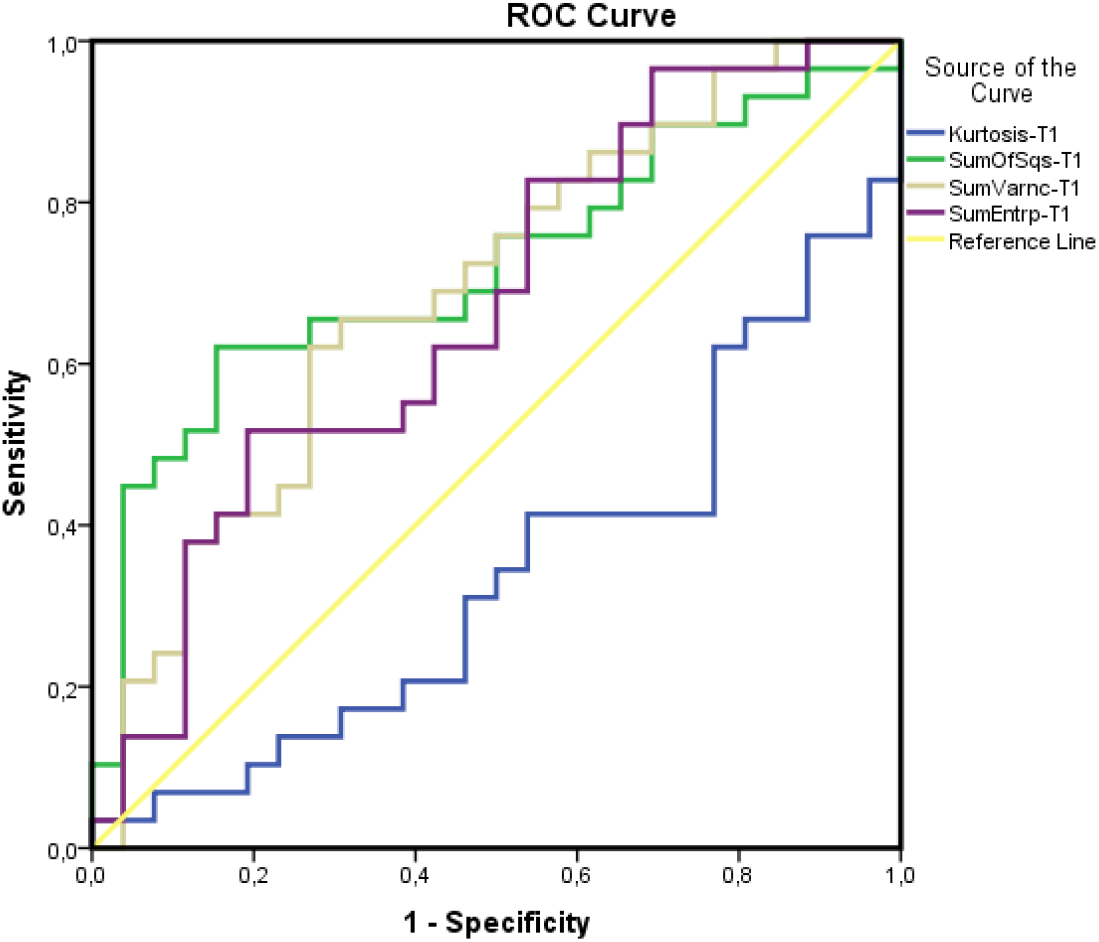
ROC analysis for T1-weighted images

Table 2: Comparison of MRI T1-weighted sequence measurements according to molecular subtypes

### Texture Analysis – T2-weighted Features

No significant differences in texture parameters were identified between subgroups for T2-weighted images.

### Morphologic MRI Features

SHH tumors were more likely to exhibit:

- nodularity or >3 lobulations (p = 0.002),
- non-midline or cerebellar hemispheric location (p < 0.001),
- heterogeneous contrast enhancement (p = 0.002),
- macrocysts >1 cm (p = 0.001), and
- peri-tumoral edema (p = 0.041).

T2 hypointensity relative to gray matter was also more common in SHH tumors. In contrast, no significant differences were observed between subgroups regarding tumor margins, hemorrhage/calcification, presence of microcysts (< 1 cm), intracranial/spinal seeding, or hydrocephalus (Figure 5).

**Figure 5.**
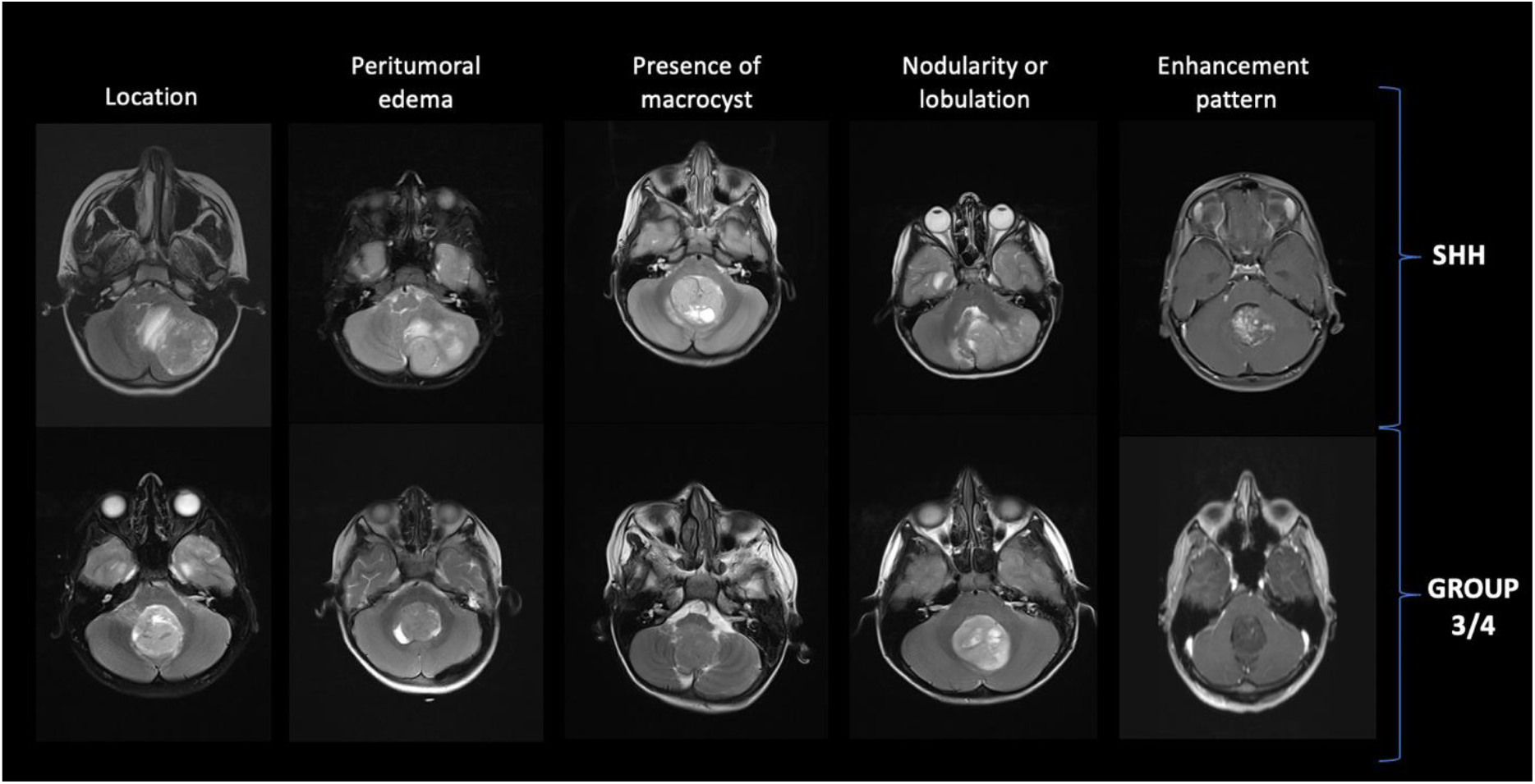
Statistically significant qualitative parameters distinguishing SHH-activated MB from Group 3/4 MB. These parameters, listed from left to right, include off-midline/cerebellar vs midline/4th ventricular location, presence or absence of peritumoral edema, macrocysts or nodularity/lobulation, and heterogeneous vs solid or none-minimal enhancement

## DISCUSSION

Medulloblastoma accounts for roughly 20–25% of pediatric brain tumors and remains the most frequent malignant tumor of the central nervous system in children [16]. It typically arises in the posterior fossa, is classified as a primitive embryonal neoplasm, and carries a WHO grade 4 designation. Over the last decade, recognition of its biological diversity has led to the definition of molecularly distinct subgroups, each with unique developmental pathways, transcriptional signatures, morphologic profiles, and clinical outcomes. According to the WHO classification, these subgroups include Wingless (WNT), Sonic Hedgehog (SHH), Group 3, and Group 4 [17].

Multiple molecular assays can distinguish these subtypes, including NanoString-based expression profiling, quantitative RT-PCR, immunohistochemical detection of specific markers, and DNA methylation profiling [18–22]. Each technique has advantages and limitations related to turnaround time, cost, and tissue requirements. In neuro-oncology imaging, conventional MRI sequences, spectroscopy, perfusion, and diffusion imaging have all been evaluated for their ability to suggest molecular subgrouping prior to surgery. Our work builds upon these approaches by incorporating texture analysis into conventional and diffusion MRI assessment.

Historically, radiological evaluation of brain tumors has focused on identifying lesion type and determining disease extent. Increasingly, it is recognized that imaging reflects biological processes such as gene expression, cellular proliferation, metabolic status, and angiogenesis [23, 24]. Radiogenomics, also termed imaging genomics, seeks to link imaging-derived phenotypes with underlying genomic or molecular features. This process involves acquiring and segmenting images, selecting and extracting features, and correlating them with molecular markers. Visual assessment of “semantic” features such as tumor volume, margins, signal intensity, enhancement characteristics, and presence of edema or necrosis remains important, but computer-based algorithms can also extract large numbers of quantitative “agnostic” features (e.g., histograms, texture metrics, wavelets) that may be more sensitive to subtle biological differences [25].

Texture analysis is one such technique, quantifying patterns of pixel intensity to reveal heterogeneity not readily visible on standard inspection. It has been applied in the imaging evaluation of tumors such as hepatocellular carcinoma, glioblastoma, and nasopharyngeal carcinoma [26–28]. Given its capacity to detect microstructural differences, we hypothesized that texture analysis could help separate MB molecular subgroups.

In our cohort, classical histology predominated among Group 3-4 tumors (73.1%), whereas SHH tumors most often exhibited desmoplastic/nodular histology (53.1%). These findings agree with Kool et al., who also found a strong correlation between SHH activation and desmoplastic/nodular subtype [3]. The medulloblastoma with extensive nodularity (MBEN) variant, detected only in SHH tumors in our series, occurred in four patients, all under one year old. While MBEN is typically reported in infants, occasional adult cases have been described [29].

Anatomically, SHH tumors originate from granule neuron precursor cells in the cerebellar external granular layer and are often hemispheric [30–33]. In our study, 62.5% of SHH cases were non-midline or hemispheric, a statistically significant distinction from Group 3-4. SHH tumors were also more frequently associated with peri-tumoral edema, consistent with prior reports noting higher edema volumes in this subgroup [7, 32, 34]. In our sample, over half of SHH tumors had measurable edema (53.1%), versus a smaller proportion in Group 3-4 (p=0.041).

The degree and pattern of contrast enhancement in SHH pathway-activated tumors have been variably reported in different studies. Dasgupta et al. found contrast enhancement in 94% of SHH subgroup medulloblastomas, with more than 50% showing a heterogeneous enhancement pattern [34]. In our study, the contrast enhancement pattern was categorized into three groups: solid (>90%), heterogeneous (10-90%), and none/minimal (<10%).

Heterogeneous contrast enhancement was detected in 58.6% of SHH-activated tumors, which was statistically significant (p=0.011). Nodularity and multi-lobulated contours (>3 lobulations) were also more common in SHH tumors, aligning with prior observations that such features may reflect multicentric cortical involvement specific to this subgroup [30, 33].

SHH subgroup medulloblastomas are frequently associated with the presence of microcysts (<1 cm) and macrocysts (>1 cm) [12]. While not classic for the SHH pathway, intratumoral macrocysts have been more commonly found in infantile SHH subgroup medulloblastomas [34]. In our study, macrocysts were found in only two of the Group 3-4 medulloblastomas, but they were present in 50% of the 32 SHH-activated medulloblastomas, which was statistically significant (p=0.001).

Diffusion characteristics likewise provided useful discriminatory power. Reddy et al. reported lower tumor-to-thalamus ADC ratios in SHH and WNT tumors compared with Group 3–4 [35]. In our dataset, the median ADC ratio was 0.75 for SHH and 0.95 for Group 3–4 tumors (p<0.001), with an optimal cut-off of 0.855 yielding 82.1% sensitivity and 92.3% specificity. The lower ADC values in SHH tumors may relate to the high density of reticulin fibers in their prevalent histologic variants, which can limit extracellular water diffusion [36–38].

It has been noted that texture analysis, a mathematical method for the quantitative analysis of variations in imaging models, shows promising diagnostic potential for various brain tumors such as glioma, meningioma, metastatic brain tumors, primary central nervous system lymphoma, and medulloblastoma [39-43]. Studies in the literature mainly focus on the use of texture analysis in differentiating brain tumors with different histology. One study demonstrated that three-dimensional texture analysis on MRI could be used to distinguish glioblastoma from primary central nervous system lymphoma [42]. Another study showed the value of various texture analysis parameters in differentiating metastatic brain tumors from high-grade gliomas [41].

In our study, ROC analysis evaluating the diagnostic value of MRI ADC measurements in distinguishing between SHH pathway-activated medulloblastoma and Group 3-4 medulloblastomas showed that the areas under the curve (AUC) for Kurtosis, Perc.01%, Perc.10%, Perc.50%, Perc.90%, and SumOfSqs ADC values were significant. These measured values were found to be weak-to-moderate in diagnostic decision-making. High Kurtosis ADC values were interpreted in favor of SHH, while low values in other measurements also supported SHH. Additionally, in the ROC curve analysis evaluating the diagnostic value of T1 measurements, the AUCs for Kurtosis, SumOfSqs, SumVarnc, and SumEntrp were found to be significant. The measured values in the T1 sequence were also found to be weak-to-moderate in diagnostic decision-making. For the SumOfSqs measurement, which had the largest AUC, the optimal cut-off point was 107.06, with a sensitivity of 62.1% and a specificity of 84.6%. Texture analysis was seen to provide valuable information in distinguishing molecular subtypes. It is possible that combinations with other MRI parameters could increase the diagnostic value, but further multi-center studies with larger patient groups are needed to support these findings.

Our study has some limitations. The cohort lacked WNT cases, and Group 3 and Group 4 tumors were analyzed together. The sample size was modest, and retrospective design may introduce selection bias. Additionally, ROI placement for quantitative measures can be subject to partial volume effects and observer variability.

## CONCLUSION

In summary, our findings indicate that combining semantic imaging features (e.g., location, cystic change, nodularity, enhancement) with quantitative metrics from diffusion and texture analysis can aid in distinguishing SHH from Group 3–4 medulloblastomas. Further, prospective multi-center studies with larger patient populations are warranted to validate these results and refine predictive models.

## Data Availability

All data produced in the present study are available upon reasonable request to the authors

